# Ethnic disparities in health-related quality of life and cognitive function after stroke in the Netherlands

**DOI:** 10.1101/2025.04.08.25325499

**Authors:** Y.X. Lee, K. Jellema, T.P.M. Vliet Vlieland, I.R. van den Wijngaard, D.E.C.M. Hofs, H.J. Arwert

## Abstract

**Background/objective:** Various studies have demonstrated, European data regarding ethnic disparities in health-related quality of life (HRQOL) after stroke is scarce and none used patient reported outcomes measurements (PROMs). This study explores whether ethnicity predict HRQOL using PROMs and cognitive function after stroke in the Netherlands.

**Patients and methods:** Patients admitted to the hospital with a first ever stroke were included. Outcome assessments included the Patient Reported Outcomes Measurement Information System (PROMIS) Profile and EuroQoL-5D (EQ-5D index and EQ-5D-L Visual Analogue Scale, VAS) for HRQOL and the PROMIS Cognitive Function for cognitive functioning. Measurements were done at hospital admission and after 3 and 12 months (EQ-5D-3L and EQ-VAS only at 12 months). Ethnicity (migration background yes/no), other demographics and stroke characteristics were collected at admission. Outcomes were compared between patients with and without a migration background by a multivariate linear mixed-effects model, adjusted for age, sex, education level and severity of stroke (NIHSS at admission).

**Results:** 262 patients were included, of whom 74 (28.2%) did and 188 (71.8%) did not have a migration background. A significant difference was observed at admission for the physical function score (estimate=3.30, SE=1.25, p=0.01), at three months for anxiety score (estimate=-2.95, SE=1.48, p=0.05) and twelve months for sleep disturbance score (estimate=-5.43, SE=1.61, p<0.01). These results are all to the disadvantage of patients with a migration background. The EQ-5D index and EQ-5D VAS at twelve months follow-up was significantly lower in patients with a migration background compared to patients without a migration background (respectively adjusted B −0.09 (95% CI −0.17; −0.01)) and adjusted B −7.27 (95% CI (−13.99; −0.56)).

**Conclusion:** Up to twelve months after hospital admission, stroke patients with a migration background had significantly worse scores regarding several domains of HRQOL compared to patients without a migration background.

## INTRODUCTION

Stroke remains the second leading cause of death and the third leading cause of death and disability combined worldwide. According to the World Stroke Organization’s Global Stroke Fact Sheet 2022, there are over 12.2 million new strokes each year globally.(1) Data from the Stroke Alliance for Europe report indicates that in 2015, there were approximately 13,789 new strokes annually in the Netherlands, with a prevalence rate of 333,8 strokes per 100,000 inhabitants.(2) In many stroke survivors, stroke is causing long-term disabilities within several domains of health, with a significant negative impact on the long-term health-related quality of life (HRQOL). This includes physical, mental and cognitive functioning and social participation.(3–6) Factors previously identified as influencing long-term stroke outcomes, aside from age, include female sex, stroke severity, comorbidity, education level, and also ethnicity.(7–9) Ethnicity is a social-political construction and is defined as a group of people that has a common national or cultural tradition(10), and its impact on various diseases, has been described before.(10,11) Ethnic disparities in end stage kidney disease in all stages of progression of the disease, the management and also the long-term quality of life were found.(12) This could also be seen in myocardial infarction.(13) These disparities may also exist in stroke patients. Previous studies indicate ethnic disparities, with patients from minority groups experiencing lower HRQOL compared to those from the predominant group in the study’s country.(14–21) This was seen on the physical component(14–16), mental component with more depressive symptoms(16) as well in the pain interference(17). However, of the aforementioned studies there are only two studies with European patients (22,28). Also, previous studies used various outcome measurements for HRQOL, but none of them used patient-reported outcome measures (PROMs). PROMs provide self-rated information about the HRQOL, independent of physicians’ interpretation and evaluation, and therefore reflect the most valuable health determinants from a patients’ perspective.(23) The International Consortium for Health Outcomes Measurement (ICHOM) has introduced a Standard Set for Stroke in 2015, including the Patient-Reported Outcomes Measurement Information System (PROMIS) Global Health. (24) The inclusion of this PROMIS measure is in line with the overall recommendation of the National Institutes of Health to use PROMIS measures in clinical care and in their funded research to enhance and standardize the use of PROMs in clinical care and research.(25) To the best of our knowledge, no studies were found that researched the role of ethnicity on post-stroke HRQOL using PROMIS Profile.

In this prospective study we aim to explore the association of ethnicity (patients with or without migration background) on HRQOL outcomes measured by PROMIS Profile among stroke survivors in a Dutch hospital population. Our secondary aim is to explore the association of ethnicity on cognitive outcomes. Our hypothesis is that a lower HRQOL and cognitive function will be found in patients with a migration background, especially on the physical component, mental component and pain interference.

## METHODS

### Study design

From September 2020 to September 2021, we included patients in a prospective cohort study at the comprehensive stroke center HMC in The Hague, a multicultural city in the Netherlands. All data were collected using questionnaires and from the patient record files. Eligible patients received the study information, an informed consent letter and the questionnaires. Written informed consent was obtained from all participants by the research nurse. The patients and, if applicable, their spouse or other caregiver, completed questionnaires about patients’ current health condition. In case of no response after two weeks, a research nurse contacted the patients by telephone to encourage them to complete the forms. Patients had the choice to withdraw from the study at any moment without reason. The study protocol was found not to fall within the scope of the Dutch Medical Research with Human Subjects Law and was approved by the institutional research board. This study was conducted according to the Declaration of Helsinki.(26) A written exemption from ethical approval was acquired from the Medical Ethics Review Committee South West Netherlands (METC nr: Z19.039). The reporting of the study was done according to The Strengthening the Reporting of Observational Studies in Epidemiology (STROBE) guidelines.(27)

### Study population

We included patients aged ≥18 years with a first ever stroke who were hospitalized. Stroke was defined as either an acute ischemic- or hemorrhagic stroke. Exclusion criteria were subarachnoid hemorrhage, traumatic intracerebral hemorrhage and venous sinus thrombosis. Patients who were unable to complete questionnaires in Dutch (e.g. due to an insufficient understanding of Dutch, severe cognitive problems or severe aphasia) were also excluded in this study.

### Measurements

Assessments included extraction of data from medical records as well as administration of an online questionnaire at admission, and 3 and 12 months thereafter. Extraction of data from the medical records was done by one researcher (Y.X. Lee). The questionnaires for sociodemographic characteristics and outcomes were collected by a research nurse.

#### Sociodemographic and stroke characteristics

##### Sociodemographic characteristics, including ethnicity

The following socio-demographic characteristics were gathered through an online questionnaire at admission: age (in years), sex (male/female), educational level was used as a proxy variable data for socioeconomic status (Low: up to and including lower technical and vocational training; Medium: up to and including secondary technical and vocational training; High: up to and including higher technical and vocational training and university), working before stroke (yes/no), smoking before admission (yes/no), alcohol utilization before admission (yes/no), having comorbidities (it was possible for patients to mark more than one answer with a choice of: transient ischemic attack/myocardial infarction/heart arrhythmia/other heart disorder/diabetes mellitus/hypertension/hypercholesterolemia/none).

##### Stroke characteristics

The stroke characteristics were extracted from the medical records; the type of stroke (ischemic/hemorrhagic), stroke side (left hemisphere/right hemisphere/other), location of the lesion (cerebrum/cerebellum/brainstem/other), acute intervention ischemic stroke (intravenous thrombolysis and/or endovascular thrombectomy or none), complications after acute intervention ischemic stroke (yes/no), severity of the stroke at admission (National Institutes of Health Stroke Scale (NIHSS), score ranging from 0 to 42, with higher scores indicating more severe neurological deficit)(28), degree of disability or dependence at admission and three months after stroke (modified Rankin Scale (mRS), a 7-point scale ranging from 0 (no symptoms) to 6 (death). A score of ≤ 2 points indicates functional independence)(29), degree of independence at admission (Barthel Index, score ranging from 0 to 20, the higher the score, the higher the level of independence),(30) length of hospitalization (in days) and discharge to home (yes/no).

##### Ethnicity

In this study, we categorized patients as a group of patients without a migration background and a group of patients with a migration background. Regarding migration background, we used the definition provided by the Dutch Central Agency for Statistics: a patient was considered to have a migration background if they were born outside the Netherlands or have at least one parent born outside the Netherlands. (31) The country of birth of patient and parents were obtained via an online questionnaire at admission.

#### Measures of HRQOL and cognitive function

The primary outcome in this study, the HRQOL, was collected and measured with two questionnaires. We used the PROMIS Profile CAT (Quest Manager, Philips Healthcare, Eindhoven, The Netherlands), by means of the CAT software of the Dutch-Flemish PROMIS Assessment Center, or the v2.1 PROMIS-29 Profile on paper. This questionnaire was completed at admission, three and twelve months after stroke onset. The PROMIS Profile CAT is a generic computer adaptive measure that assesses seven important HRQOL domains using the equivalent item banks: Physical Function v1.2, Anxiety v1.0, Depression v1.0, Fatigue v1.0, Sleep Disturbance v1.0, Ability to Participate in Social Roles and Activities v2.0, Pain Interference v1.1 and pain intensity (NRS, scores 0-10). Furthermore, for the cognitive function outcome, the PROMIS Cognitive function CAT or short form was applied. Based on the underlying Item Response Theory (IRT) model (using the standard USA item parameters), highly informative subsets of items were selected from the corresponding item banks to provide optimal measurement properties.

The versions of the PROMIS Profile that are composed of short forms, consist of a total of 29, 43 or 57 items. Patients unable to fill in the CAT version used the PROMIS 29 and the PROMIS Cognitive Function on paper. Scores of each CAT were presented as an IRT based T-score. A T-score of 50 is the average for the USA general population with a standard deviation (SD) of 10. A higher score annotates more of the specific construct. A difference of 3 T-score points is considered a relevant change.(32) For the NRS score of pain an MID (minimal important difference) of 2 points was considered.(33) The EuroQol-5D 3L (EQ-5D) was also administered at twelve months after stroke. The EQ-5D is a standardized generic questionnaire that scores five health domains (mobility, self-care, daily activities, pain/discomfort and anxiety/depression) to assess HRQOL. The five questions about the current health status have each three levels (no problems, some problems, extreme problems/unable to).(34) A Dutch formula attaches a weight to each level, and can be combined into a single utility value (EQ-5D index) with a value of 1.00 representing “full health” and a value lower than zero representing “worse than death”.(35–37) The second part of the EQ-5D consist of a visual analog scale (EQ-5D VAS) and provides an overall assessment of the current health state with a score from 0 (the worst health you can imagine) to 100 (the best health you can imagine).(38) There is an observational longitudinal survey published about the MCID of EQ-5D index of stroke patients.(39) They found an MID ranging from 0.08 to 0.12 by using anchor-based approaches. One study showed an MID ranging from 8.61 to 10.82 for the EQ-5D VAS in stroke patients.(40)

### Data analysis

All analyses were conducted using R version 4.4.3 (R Foundation for Statistical Computing, Vienna, Austria), except for the descriptive statistics, EQ 5D index and EQ 5D VAS. These were analyzed using SPSS Statistics, version 27 (Armonk, NY: IBM Corp). Descriptive statistics were calculated for baseline characteristics. Continuous variables were presented as means with standard deviations (SD), and categorical variables as numbers with percentages. The characteristics of patients with or without a migration background were compared using unpaired t-tests for continuous variables and Chi-Square tests for categorical variables. A multivariate linear regression analysis was conducted to evaluate the impact of possible confounders (age at admission, sex, education level and severity of stroke using the NIHSS at admission). To examine changes in PROMIS outcome measures over time (baseline, 3 months and 12 months) and the effect of ethnicity, a multivariate linear mixed-effects model was used, applying the lme4 package. The model included a random intercept and a random slope for time per individual to account for within-subject correlations over time. Fixed effects included time, ethnicity, and their interaction term (ethnicity × time). Age at admission, sex, education level, and stroke severity (NIHSS at admission) were included as covariates. The model estimates were reported as crude and adjusted unstandardized beta coefficients (B) with 95% confidence intervals (CI). To further explore differences between ethnicity at the three different time points, post hoc pairwise comparisons with Bonferroni correction were performed. Since the model with a random intercept and random slope is too complicated to perform a post-hoc analysis, the model was reduced to a model with only a random intercept.(41–43) Statistical significance was set at a p-value of ≤ 0.05. Missing values on the outcome variable at one or more time points were not imputed. However, linear mixed models allow for the inclusion of individuals with partially missing outcome data, as long as at least one observation is available. This approach assumes that data are missing at random.

## RESULTS

A total of 475 patients were considered eligible for participation. 263 patients provided informed consent. We excluded one patient due to lack of information about ethnicity. We observed significant differences in sex, age and NIHSS at admission between the included and not-included patients. The included patients were younger, included more men and had a higher NIHSS at admission compared to the not-included patients (Table 1). Patients received the EQ-5D questionnaire and VAS question twelve months after stroke, PROMIS Profile and PROMIS Cognitive Function at admission, three months and twelve months after stroke.

**TABLE 1.** DIFFERENCES BASELINE CHARACTERISTICS IN- AND EXCLUDED PATIENTS.

|  | **INCLUDED PATIENTS**<br>**N=262** | **EXCLUDED PATIENTS**<br>**N=213** | **P VALUE** |
| --- | --- | --- | --- |
| Median age, years (IOR) | 71.00 (18) | 75.00 (21) | <u>**0.048**</u> |
| Sex, male – n (%) | 151 (57.6) | 102 (47.9) | <u>**0.042**</u> |
| NIHSS at admission – median (IOR) | 4.00 (4) | 9.00 (12) | <u>**<0.001**</u> |

IOR=Interquartile Range, NIHSS = National Institutes of Health Stroke Scale

In Table 2 we present the patient characteristics of the patients with and without a migration background. Patients with a migration background were significantly younger compared to patients without a migration background, respectively median age of 68.00 (IOR 18) and 73.00 (IOR 16) years; p=0.01; no differences were observed regarding sex, educational level and stroke severity (NIHSS score). Patients with a migration background received less acute reperfusion therapy (intravenous thrombolysis and/or endovascular vascular therapy) in case of an ischemic stroke compared to patients without a migration background (29.7% vs 44.7%; p=0.04) and patients with a migration background had a longer length of hospital stay compared to patients without a migration background (6.42 days (5.35 SD) vs 4.99 days (4.88 SD); p=0.04).

**TABLE 2.**
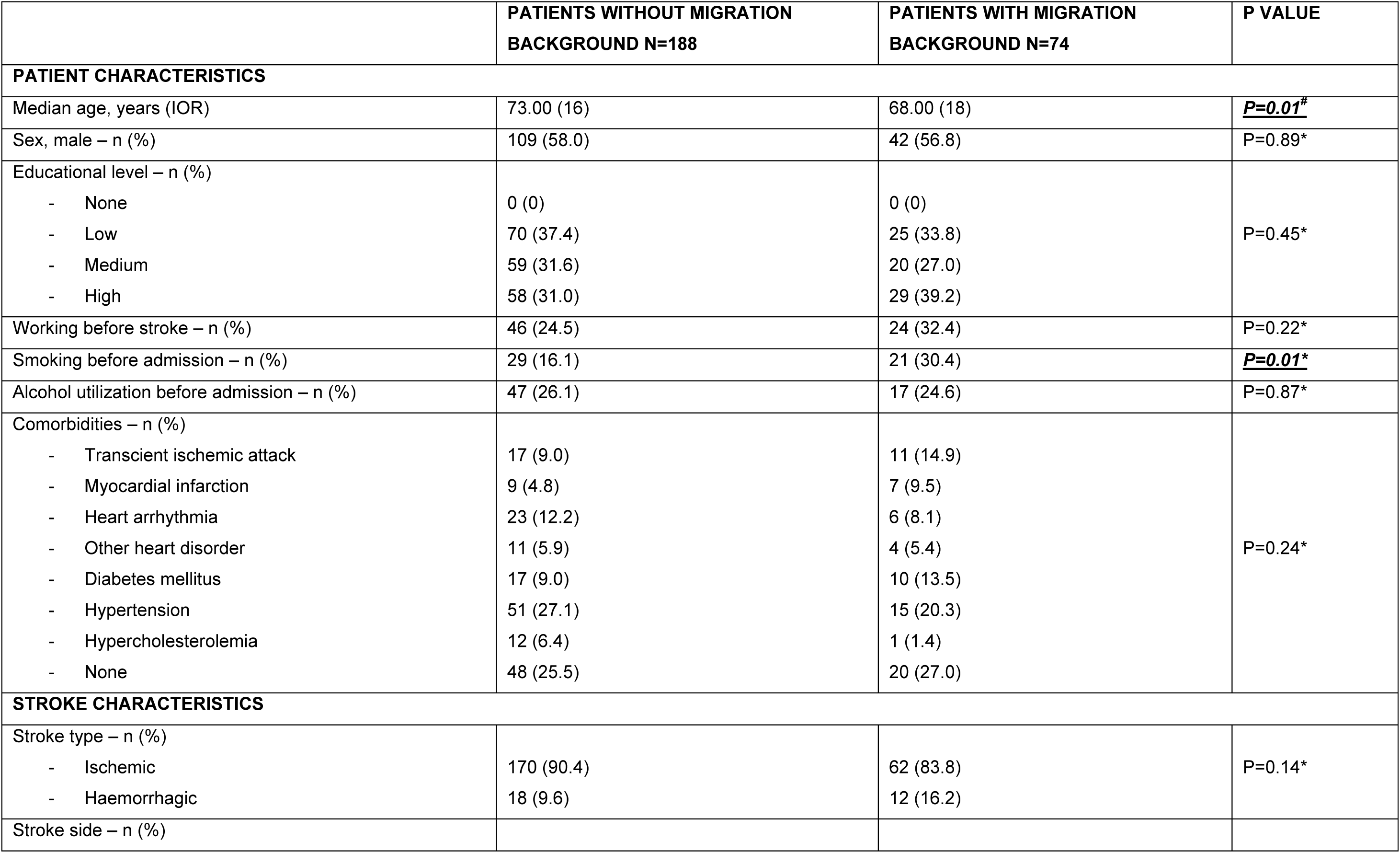
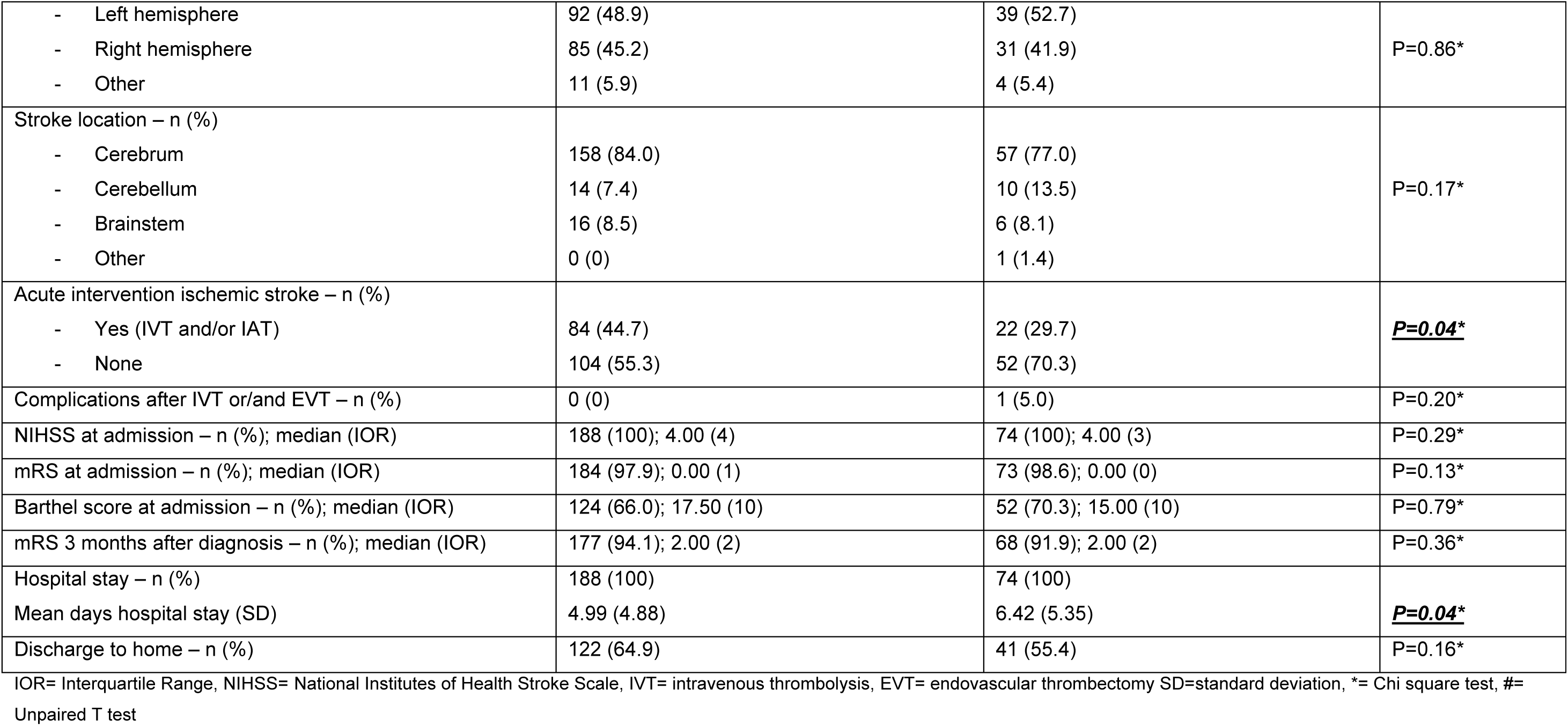
BASELINE CHARACTERISTICS INCLUDED PATIENTS.

The results of all eight domains of the PROMIS Profile CAT, the PROMIS Cognitive function the EQ-5D index and EQ-5D VAS are presented in Table 3. All scores of the PROMIS Profile CAT (with the exception of the anxiety- and sleep disturbance scores) and the PROMIS Cognitive function scores in- or decreased significantly over time. An increasing physical function score and ability to participate in social roles and activities score over time (respectively B= 7.08, 95% CI: 6.21 to 7.95; and B= 11.18, 95% CI:9.91 to 12.46) means a better outcome over time. An increasing depression-(B= 1.30, 95% CI: 0.45 to 2.15), fatigue-(B= 1.33, 95% CI: 0.49 to 2.18), pain interference-(B= 2.17, 95% CI: 1.00 to 3.34) and pain intensity score (B= 0.40, 95% CI: 0.12 to 0.69) means a worse outcome over time. The cognitive function score (B= −3.23, 95% CI: −4.14 to −2.32) decreased over time meaning a worse outcome over time. There were significantly differences on the physical function-, anxiety-, and sleep disturbance score between patients without and with a migration background. For the physical function score, we found significantly lower scores in patients with a migration background compared to patients without a migration background (B= −5.60, 95% CI: −9.01 to −2.19). A significant interaction between ethnicity and time was observed (B= 1.87, 95% CI: 0.15 to 3.60), indicating a different trajectory in physical function over time between ethnic groups. To explore this interaction further, post hoc pairwise comparisons between both groups were conducted at each time point using Bonferroni correction for multiple testing. At admission, physical function scores were significantly higher in the group without a migration background compared to the group with a migration background (estimate = 3.30, SE= 1.25, p=0.01). At 3 months, this difference was no longer statistically significant (estimate = 2.54, SE= 1.38, p=0.07). At 12 months, no difference was observed between groups (estimate = –0.19, SE= 1.41, p=0.90). For the anxiety score we found a significant interaction between ethnicity and time (B= 1.64, 95% CI: 0.07 to 3.20=), indicating a different trajectory in anxiety over time between groups. To explore this interaction further, post hoc pairwise comparisons between both groups were conducted at each time point using Bonferroni correction for multiple testing. At admission, there was no significant difference between the two groups (estimate = 0.29, SE= 1.32, p=0.83). At 3 months, anxiety scores were significantly higher in the group with a migration background compared to the group without a migration background (estimate = −2.95, SE= 1.48, p=0.05). At 12 months, this difference was no longer statistically significant (estimate = −2.72, SE= 1.53, p=0.08). For the sleep disturbance scores, there was a significant interaction between ethnicity and time (B= 2.17, 95% CI: 0.61 to 3.73), indicating a different trajectory in sleep disturbance over time between groups. To explore this interaction further, post hoc pairwise comparisons between groups were conducted at each time point using Bonferroni correction for multiple testing. At admission and at 3 months, there were no significant differences between the two groups (respectively estimate = −1.01, SE= 1.39, p=0.47 and estimate = −2.15, SE= 1.56, p=0.17). At 12 months, sleep disturbance scores were significantly higher in the group with a migration background compared to the group without a migration background (estimate = −5.43, SE= 1.61, p<0.01). There was no significant effect of ethnicity (B= −3.36, 95% CI: −6.83 to −0.11, p=0.06) on the cognitive function scores. However, the p-value is very close to significance. In addition, there was no interaction between time and ethnicity (B= 0.56, 95% CI: −1.25 to 2.36, p=0.54). The significant difference for physical function was the only one with a difference of >3 T-score points. No significant differences were found between patients without and with a migration background for the depression-, fatigue-, ability to participate in social roles and activities-, pain interference- and pain scores.

**TABLE 3.** RESULTS PATIENTS WITH OR WITHOUT MIGRATION BACKGROUND AND PROMS AT ADMISSION, 3 MONTHS AND 12 MONTHS AFTER STROKE; AND EQ-5D INDEX AND EQ-5D VAS 12 MONTHS AFTER STROKE.

|  | PMB-<br>N (%) | PMB-<br>Mean (lower-upper CL) | PMB+<br>N (%) | PMB+<br>Mean (lower-upper CL) | B COEFFICIENT<br>(95% CI)<sup>#</sup> | ESTIMATE*, SE<br>(p-value) |
| --- | --- | --- | --- | --- | --- | --- |
| **PROMIS Profile CAT Domain** |  |  |  |  |  |  |
| Physical function |  |  |  |  |  |  |
| - At admission | 170 (90.4) | 29.2 (27.9-30.5) | 62 (83.8) | 25.9 (23.8-28.0) |  | **<u>3.30, 1.25 (0.01)</u>** |
| - 3 months | 143 (76.3) | 42.3 (40.9-43.7) | 47 (64.5) | 39.8 (37.4-42.1) |  | 2.54, 1.38 (0.07) |
| - 12 months | 134 (71.6) | 43.0 (41.6-44.4) | 44 (57.9) | 43.2 (40.8-45.6) |  | -0.19, 1.41, (0.90) |
| - Time (as predictor) |  |  |  |  | **<u>7.08 (6.21; 7.95)</u>** |  |
| - Ethnicity (as predictor) |  |  |  |  | **<u>-5.60 (-9.01; -2.19)</u>** |  |
| - Interaction ethnicity and time |  |  |  |  | **<u>1.87 (0.15; 3.60)</u>** |  |
| Anxiety |  |  |  |  |  |  |
| - At admission | 172 (91.5) | 51.4 (50.0- 52.8) | 67 (90.5) | 51.1 (48.9-53.3) |  | 0.29, 1.32 (0.83) |
| - 3 months | 135 (71.8) | 53.0 (51.4-54.5) | 49 (66.2) | 55.9 (53.4-58.4) |  | **<u>-2.95, 1.48 (0.05)</u>** |
| - 12 months | 128 (68.1) | 52.7 (51.2-54.3) | 44 (59.5) | 55.5 (52.9-58.0) |  | **<u>-2.72, 1.53 (0.08)</u>** |
| - Time (as predictor) |  |  |  |  | 0.73 (-0.07; 1.53) |  |
| - Ethnicity (as predictor) |  |  |  |  | -1.49 (-4.90; 1.93) |  |
| - Interaction ethnicity and time |  |  |  |  | **<u>1.64 (0.07; 3.20)</u>** |  |
| Depression |  |  |  |  |  |  |
| - At admission | 150 (79.8) | 49.0 (47.6-50.4) | 62 (83.8) | 49.9 (47.8-52.1) |  | -0.92, 1.32 (0.49) |
| - 3 months | 125 (66.5) | 51.8 (50.3-53.3) | 45 (60.8) | 52.9 (50.5-55.4) |  | -1.15, 1.48 (0.44) |
| - 12 months | 118 (62.8) | 51.5 (50.0-53.0) | 37 (50.0) | 54.4 (51.7-57.1) |  | -2.93, 1.57 (0.06) |
| - Time (as predictor) |  |  |  |  | **<u>1.30 (0.45; 2.15)</u>** |  |
| - Ethnicity (as predictor) |  |  |  |  | -0.32 (-3.72; 3.09) |  |
| - Interaction ethnicity and time |  |  |  |  | 1.01 (-0.68; 2.70) |  |
| Fatigue |  |  |  |  |  |  |
| - At admission | 177 (94.1) | 48.3 (46.7-49.8) | 68 (91.9) | 49.1 (46.6-51.6) |  | -0.85, 1.50 (0.57) |
| - 3 months<br>- 12 months<br>- Time (as predictor)<br>- Ethnicity (as predictor)<br>- Interaction ethnicity and time | 141 (75.0)<br>134 (71.3) | 51.3 (49.6-52.9)<br>50.8 (49.1-52.5) | 47 (63.5)<br>43 (58.1) | 54.9 (52.0-57.8)<br>53.3 (50.3-56.2) | **<u>1.33 (0.49; 2.18)</u>**<br>0.24 (-3.71; 4.18)<br>0.99 (-0.69; 2.67) | **<u>-3.65, 1.70 (0.03)</u>**<br>-2.45, 1.74 (0.16) |
| Sleep disturbance<br>- At admission<br>- 3 months<br>- 12 months<br>- Time (as predictor)<br>- Ethnicity (as predictor)<br>- Interaction ethnicity and time | 169 (89.9)<br>137 (72.9)<br>125 (66.5) | 48.7 (47.3-50.2)<br>49.0 (47.4-50.6)<br>47.4 (45.7-49.0) | 67 (90.5)<br>47 (63.5)<br>43 (58.1) | 49.7 (47.4-52.0)<br>51.2 (48.5-53.8)<br>52.8 (50.1-55.5) | -0.64 (-1.43; 0.15)<br>-1.44 (-5.09; 2.20)<br>**<u>2.17 (0.61; 3.73)</u>** | -1.01, 1.39 (0.47)<br>-2.15, 1.56, (0.17)<br>**<u>-5.43, 1.61 (<0.01)</u>** |
| Ability to participate in social roles and activities<br>- At admission<br>- 3 months<br>- 12 months<br>- Time (as predictor)<br>- Ethnicity (as predictor)<br>- Interaction ethnicity and time | 173 (92.0)<br>132 (70.2)<br>131 (69.7) | 29.4 (27.8-31.1)<br>49.9 (48.1-51.8)<br>51.0 (49.1-52.9) | 66 (89.2)<br>45 (60.8)<br>42 (56.8) | 29.5 (26.9-32.2)<br>45.8 (42.6-49.0)<br>50.5 (47.2-53.9) | **<u>11.18 (9.91; 12.46)</u>**<br>-0.84 (-6.05; 4.37)<br>-0.28 (-2.80; 2.24) | -0.10, 1.61 (0.95)<br>**<u>4.16, 1.91 (0.03)</u>**<br>0.46, 1.96 (0.81) |
| Pain interference<br>- At admission<br>- 3 months<br>- 12 months<br>- Time (as predictor)<br>- Ethnicity (as predictor)<br>- Interaction ethnicity and time | 92 (48.9)<br>102 (54.3)<br>90 (47.9) | 49.5 (47.7-51.4)<br>54.7 (52.9-56.5)<br>54.1 (52.2-55.9) | 45 (60.8)<br>36 (48.6)<br>34 (45.9) | 51.0 (48.3-53.6)<br>55.6 (52.7-58.6)<br>55.0 (52.0-58.0) | **<u>2.17 (1.00; 3.34)</u>**<br>1.11 (-3.55; 5.78)<br>-0.08 (-2.22; 2.07) | -1.45, 1.65 (0.38)<br>-0.90, 1.75 (0.61)<br>-0.90, 1.81 (0.62) |
| Pain intensity (NRS)<br>- At admission | 180 (95.7) | 1.73 (1.30-2.16) | 69 (93.2) | 2.22 (1.53-2.91) |  | -0.49, 0.41 (0.24) |
| - 3 months | 144 (76.6) | 2.53 (2.06-3.00) | 47 (63.5) | 3.40 (2.59-4.22) |  | -0.87, 0.48 (0.07) |
| - 12 months | 102 (54.3) | 2.43 (1.89-2.97) | 32 (43.2) | 3.17 (2.21-4.13) |  | -0.74, 0.56 (0.19) |
| - Time (as predictor) |  |  |  |  | **<u>0.40 (0.12; 0.69)</u>** |  |
| - Ethnicity (as predictor) |  |  |  |  | 0.33 (-0.84; 1.50) |  |
| - Interaction ethnicity and time |  |  |  |  | 0.18 (-0.40; 0.76) |  |
| **PROMIS Cognitive function** |  |  |  |  |  |  |
| - At admission | 179 (95.2) | 55.0 (53.5-56.5) | 69 (93.2) | 52.4 (50.0-54.7) |  | 2.65, 1.43 (0.06) |
| - 3 months | 135 (71.8) | 49.6 (47.9-51.2) | 42 (56.8) | 46.6 (43.7-49.4) |  | 3.00, 1.69 (0.08) |
| - 12 months | 129 (68.6) | 48.9 (47.2-50.6) | 41 (55.4) | 47.6 (44.7-50.5) |  | 1.29, 1.70 (0.45) |
| - Time (as predictor) |  |  |  |  | **<u>-3.23 (-4.14; -2.32)</u>** |  |
| - Ethnicity (as predictor) |  |  |  |  | -3.36 (-6.83; -0.11) |  |
| - Interaction ethnicity and time |  |  |  |  | 0.56 (-1.25; 2.36) |  |
| **EQ-5D index at 12 months** | 125 (67.0) | 0.797 (0.22) | 40 (55.4) | 0.727 (0.26) | **<u>-0.09 (-0.17; -0.01)</u>** |  |
| **EQ-5D VAS at 12 months** | 126 (67.0) | 73.67 (18.15) | 41 (55.4) | 67.22 (20.66) | **<u>-7.27 (-13.99; -0.56)</u>** |  |

PMB- = patients without migration background, PMB+ = patients with migration background, NRS = Numeric rating scale, SD = standard deviation, CL = confidence level, CI = confidence interval, B = unstandardized beta, \* = estimated contrast between PMB- minus PMB+, SE = standard error, # = corrected for age, sex, educational level and NIHSS at admission

After adjusting for possible confounders, the mean EQ-5D index as well as the mean EQ-5D VAS showed significantly lower scores in patients with a migration background, with respectively a difference of −0.09 (95% CI: −0.17 to −0.01) and −7.27 (95% CI: −13.99 to −0.56) points. The significant difference in EQ-5D index was also relevant according the aforementioned MID higher than 0.08. The difference in EQ-5D VAS was not relevant with an MID lower than 8.61.

## DISCUSSION

In this Dutch stroke population, we found that patients with a migration background who had a first stroke, have a significant and relevant lower quality of life compared to patients without a migration background. This difference was present after adjustment for possible confounders (age, sex, educational level and stroke severity). This disparity was observed at admission at three- and twelve months poststroke. The lower quality of life in these patients is not only determined by a poorer physical health but also by a worse mental health (higher anxiety score) and more sleep disturbance. However, only the difference in physical function score was considered relevant. In addition, patients with a migration background also reported worse outcomes on the EQ-5D index and EQ-5D VAS twelve months after stroke.

European data according this topic is limited, even more for data about the Dutch population. Our findings align with a systematic review from 2024 that included eleven studies (four prospective and seven retrospective studies), compromising a total of 12430 patients.(44) Ten of these studies identified ethnic disparities in quality of life, in which eight studies reported a lower HRQOL in patients from a minority ethnic group compared to the predominant ethnic group in that particularly country. However, included studies had a large heterogeneity in study population and outcome measurements. Given the differences in healthcare access across countries, caution is needed when comparing results from different countries. Notably, in our previous retrospective study we found a lower HRQOL in patients with a non-European background compared to patients with a European background, along with a higher level of depressive symptoms and more frequent physician visits, persisting even five years after stroke.(16) Other potential explanations for the differences in HRQOL between patients with and without a migration background may include prestroke differences in HRQOL. A previous study analyzed differences in HRQOL among elderly immigrants of different ethnic groups in general society in the Netherlands, including Moroccans, Turks and Moluccans (Indonesia). While this study found HRQOL disparities, the differences were primarily affected by other factors, such as multimorbidity and loneliness, not ethnicity.(45)

Therefore, our finding of a lower poststroke HRQOL are unlikely the result of a possible lower prestroke HRQOL in patients with a migration background. Another explanation of ethnical inequalities in HRQOL consists of differences in socioeconomic factors. A lower income can be an independent negative predictor for HRQOL in poststroke patients.(46) Data regarding educational level served as a proxy variable in this respect, and was corrected for in the regression analysis.

A strength of this study is the multi-ethnic cohort, along with the inclusion of both EQ-5D and PROMIS questionnaires, which capture a broad range of HRQOL aspects relevant to patients’ experiences. Ethnicity was defined in an objective way by reporting the country of birth of the patients and their parents.

A possible explanation for the differences between patients with and without a migration background could also be a disparity in acute stroke treatment between groups. We found a lower percentage of acute reperfusion therapy for ischemic stroke patients with a migration background compared to patients without a migration background. Acute reperfusion therapy (intravenous thrombolysis or endovascular therapy) has proven to improve mortality rates and functional outcomes.(47,48) Whether a higher percentage of late presentations of stroke or more misdiagnoses in patients with a migration background compared to patients without a migration background played a role is not yet known.

There are also several limitations to take into account. There were significant differences in sex, age and severity of stroke between included and excluded patients. This may be due to patients with more severe strokes being less likely to be able to complete questionnaires (for example due to severe cognitive problems or severe aphasia). Therefore, our findings should be interpreted within the context of this selected group and may not be generalizable to older patients or those with more severe strokes.

A higher incidence of stroke with a lower mean age in blacks and Hispanics compared to whites has been demonstrated in earlier studies.(49,50) This is in line with our patient group with a migration background with a significant lower mean age compared to patients without a migration background. The ethnic disparity in incidence was largely explained by socio-economic status (insurance status and educational level), however persisted after correction for these factors.(49) There is also an increased burden of vascular disease risk factors in blacks and Hispanics(49,50), such as smoking, which is also seen to be significant higher in our patients with migration background compared to the patients without a migration background. Furthermore, while we corrected for several possible confounders, it is possible that other unmeasured factors, such as genetics, may have influenced the outcomes. Finally, the study design may introduce a risk of selection bias due to loss to follow-up.

In conclusion, this study shows that disparities exist regarding quality of life in Dutch stroke survivors, in which patients with a migration background scored significantly and relevantly lower on different aspects of quality of life. Ethnicity seems to be an independent factor of influence on poststroke HRQOL. Identifying specific patient categories with higher risks of residual complaints and limitations after stroke is important for the improvement of poststroke healthcare in these specific groups, in the context of value-based healthcare. More inclusive data on poststroke outcomes across different ethnic populations is needed to provide a fuller and more representative understanding of poststroke outcomes in diverse population groups. Future studies should focus on the underlying mechanisms of ethnic disparities in HRQOL and examine whether healthcare utilization indeed differs across these groups. Such insights would be valuable for developing patient care pathways that target high risk groups, aiming to achieve equal HRQOL outcomes for all patients after stroke.

## Data Availability

Data can be given upon request

## Statistical analysis completed

D.E.C.M. Hofs, H.J. Arwert, K. Jellema, Y.X. Lee There are no “supplementary data” to this manuscript.

## Author contributions

Drafting/revising the manuscript for content, including medical writing for content: H.J. Arwert, K. Jellema, Y.X. Lee, T.P.M. Vliet Vlieland, I.R. van den Wijngaard

Study concept or design: H.J. Arwert, K. Jellema, Y.X. Lee, I.R. van den Wijngaard

Analysis or interpretation of data: H.J. Arwert, D.E.C.M. Hofs, K. Jellema, Y.X. Lee

